# Severe motor impairment is associated with lower contralesional brain age in chronic stroke

**DOI:** 10.1101/2024.10.26.24316190

**Authors:** Gilsoon Park, Mahir H. Khan, Justin W. Andrushko, Nerisa Banaj, Michael R. Borich, Lara A. Boyd, Amy Brodtmann, Truman R. Brown, Cathrin M. Buetefisch, Adriana B. Conforto, Steven C. Cramer, Michael Dimyan, Martin Domin, Miranda R. Donnelly, Natalia Egorova-Brumley, Elsa R. Ermer, Wuwei Feng, Fatemeh Geranmayeh, Colleen A. Hanlon, Brenton Hordacre, Neda Jahanshad, Steven A. Kautz, Mohamed Salah Khlif, Jingchun Liu, Martin Lotze, Bradley J. MacIntosh, Feroze B. Mohamed, Jan E. Nordvik, Fabrizio Piras, Kate P. Revill, Andrew D. Robertson, Christian Schranz, Nicolas Schweighofer, Na Jin Seo, Surjo R. Soekadar, Shraddha Srivastava, Bethany P. Tavenner, Gregory T. Thielman, Sophia I. Thomopoulos, Daniela Vecchio, Emilio Werden, Lars T. Westlye, Carolee J. Winstein, George F. Wittenberg, Chunshui Yu, Paul M. Thompson, Sook-Lei Liew, Hosung Kim

## Abstract

**Background:** Stroke leads to complex chronic structural and functional brain changes that specifically affect motor outcomes. The brain-predicted age difference (brain-PAD) has emerged as a sensitive biomarker. Our previous study showed higher global brain-PAD associated with poorer motor function post-stroke. However, the relationship between local stroke lesion load, regional brain age, and motor impairment remains unclear.

**Methods:** We studied 501 individuals with chronic unilateral stroke (>180 days post-stroke) from the ENIGMA Stroke Recovery Working Group dataset (34 cohorts). Structural T1-weighted MRI scans were used to estimate regional brain-PAD in 18 predefined functional subregions via a graph convolutional network algorithm. Lesion load for each region was calculated based on lesion overlap. Linear mixed-effects models assessed associations between lesion size, local lesion load, and regional brain-PAD. Machine learning classifiers predicted motor outcomes using lesion loads and regional brain-PADs. Structural equation modeling examined directional relationships among corticospinal tract lesion load (CST-LL), ipsilesional brain-PAD, motor outcomes, and contralesional brain-PAD.

**Findings:** Larger total lesion size was positively associated with higher ipsilesional regional brain-PADs (older brain age) across most regions (p□<□0.05), and with lower contralesional brain-PAD, notably in the ventral attention-language network (p□<□0.05). Higher local lesion loads showed similar patterns. Specifically, lesion load in the salience network significantly influenced regional brain-PADs across both hemispheres. Machine learning models identified CST-LL, salience network lesion load, and regional brain-PAD in the contralesional frontoparietal network as the top three predictors of motor outcomes. Structural equation modeling revealed that larger stroke damage was associated with poorer motor outcomes (β□=□-0.355, p□<□0.001), which were further linked to younger contralesional brain age (β□=□0.204, p□<□0.001), suggesting that severe motor impairment is linked to compensatory decreases in contralesional brain age.

**Interpretation:** Our findings reveal that larger stroke lesions are associated with accelerated aging in the ipsilesional hemisphere and paradoxically decelerated brain aging in the contralesional hemisphere, suggesting compensatory neural mechanisms. Assessing regional brain age may serve as a biomarker for neuroplasticity and inform targeted interventions to enhance motor recovery after stroke.

**Fundings:** Micheal J Fox Foundation, National Institutes of Health, Canadian Institutes of Health Research, National Health and Medical Research Council, Australian Brain Foundation, Wicking Trust, Collie Trust, and Sidney and Fiona Myer Family Foundation, National Heart Foundation, Hospital Israelita Albert Einstein, Australian Research Council Future Fellowship, Wellcome Trust, National Institute for Health Research Imperial Biomedical Research Centre, European Research Council, Deutsche Forschungsgemeinschaft, REACT Pilot, National Resource Center, Research Council of Norway, South-Eastern Norway Regional Health Authority, Norwegian Extra Foundation for Health and Rehabilitation, Sunnaas Rehabilitation Hospital HT, University of Oslo, and VA Rehabilitation Research and Development

## Introduction

Stroke induces complex and distinct structural and functional changes in the brain different brain regions and networks.^1^ Total and regional brain volumes rapidly decline within the first year post-stroke, beyond the atrophy rates observed in normal aging.^2^ Disruptions to white matter integrity, especially in the corticospinal tract and corpus callosum,^3,4^ strongly predict motor impairment at chronic timepoints. Moreover, patients with stroke often exhibit increased complexity in functional connectivity compared to healthy controls, suggesting compensatory mechanisms and neural reorganization to overcome damaged connections.^5^ Importantly, in chronic stroke, brain changes may result from lesion-induced damage or use-dependent reorganization supporting adaptive behaviors.^6^ Quantifying regional variations in these changes could identify precise neural biomarkers or therapeutic targets to enhance rehabilitation and understand neuroplasticity after stroke.

Brain age estimation, based on neuroimaging features, offers a promising approach to quantify regional structural changes of post-stroke.^7,8^ The brain-predicted age difference (brain-PAD)—the difference between predicted brain age and chronological age—has emerged as a sensitive biomarker, with higher global brain-PAD associated with poorer post-stroke motor outcomes.^9^ However, global brain age lacks specificity. Investigating regional brain age may provide more granular insights into patterns of secondary stroke damage across the ipsilesional and contralesional hemispheres. This approach could reveal how undamaged brain areas compensate for damaged regions, identifying neural targets for interventions aimed at slowing accelerated aging processes and improving recovery, such as targeted drug treatments or non-invasive brain stimulation for personalized stroke rehabilitation strategies, particularly in chronic stroke.^10^

In this study, we examined the relationship between focal stroke damage, motor outcomes, and regional brain age in individuals with chronic stroke. We employed a graph convolutional network (GCN) algorithm to estimate regional brain-PAD using cortical features of predefined functional subregions. We hypothesized that higher focal lesion damage would be strongly associated with older regional brain ages in the ipsilesional hemisphere and worse functional outcomes. Additionally, previous literature has demonstrated greater contralesional hemisphere activity in severe stroke cases.^11,12,13^ Thus, we hypothesized that both higher ipsilesional regional brain-PAD and lower contralesional regional brain-PAD would be important features in predicting better motor outcomes. Finally, to assess directional relationships, we performed mediation analysis to examine whether and how ipsilesional brain age may mediate the relationship between lesion damage and motor outcomes, and, subsequently, how motor outcomes may impact contralesional brain age.

## Materials and Methods

### Participants

We analyzed data from two sets of participants for algorithm development and analysis, respectively.

The regional brain age prediction algorithm, as described previously,^14^ was trained on a subset of data from the UK Biobank database, a large-scale biomedical repository containing genetic, imaging, and health information from over 500,000 individuals in the United Kingdom.^15^ Briefly, we included participants with MRI brain imaging who were non-Hispanic white and excluded any subjects with self-reported or hospital-recorded history of any neurological disorders, resulting in 17,791 individuals (52.7% female, mean age 63.2 ± 7.4 years) for algorithm development.

We then used the ENIGMA Stroke Recovery Working Group dataset of individuals with stroke, a multi-site repository containing retrospective studies of neuroimaging data, demographic information, and functional outcome measures.^16^ Data were frozen for this analysis on June 23, 2023. We included any subjects who had a motor outcome measure who were defined to be in the chronic stage of stroke recovery (180 days or more since stroke),^17^ had a chronological age of 45 years or older, had the presence of lesion in only one hemisphere (i.e., unilateral lesion), and who had a successful extraction of cortical thickness (Fig. 1).

**Figure 1.**
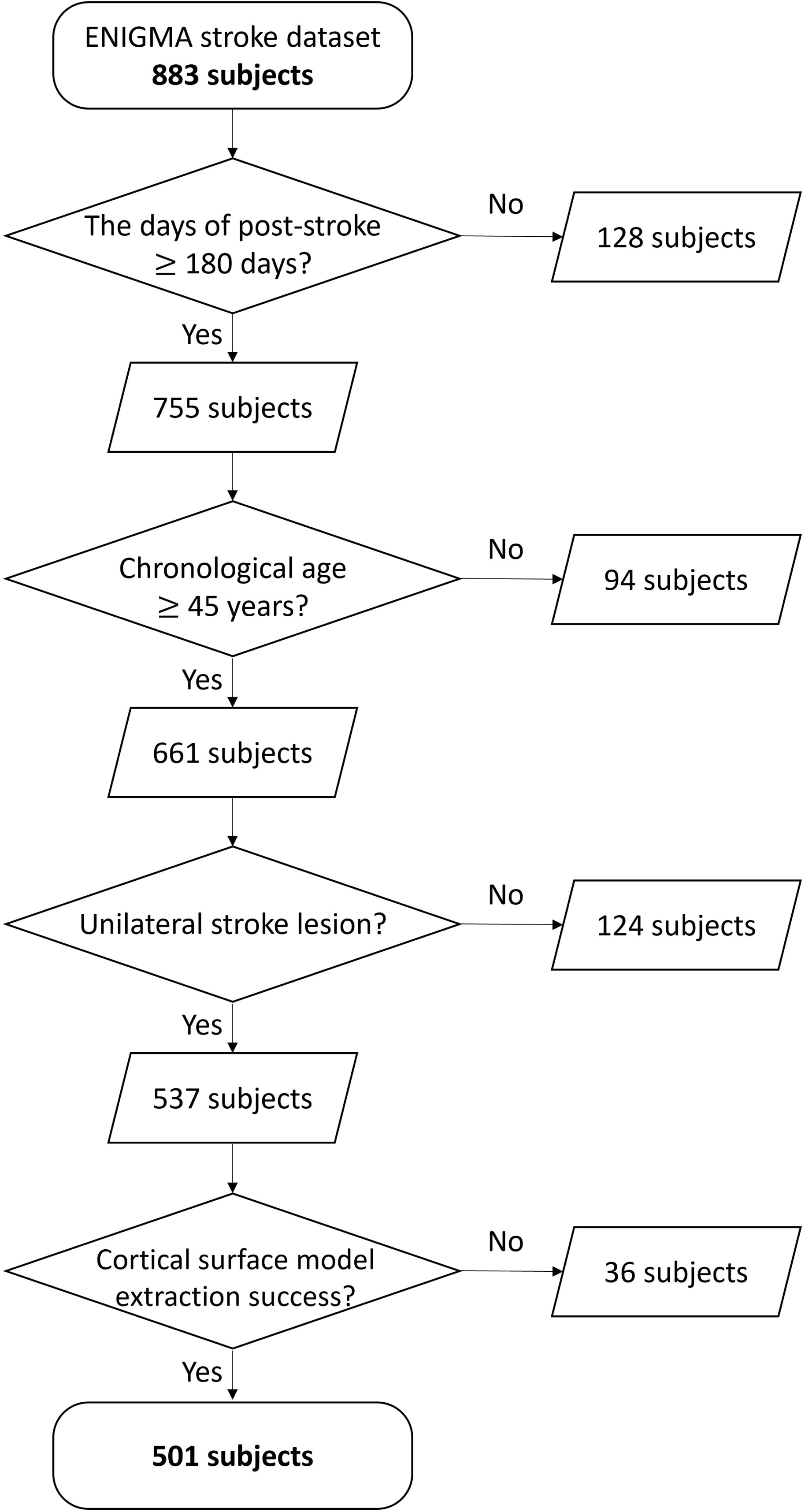
The data selection flowchart for stroke subjects. From the 883 initial subjects, we selected 501 participants who met the following four criteria: (1) post-stroke duration of at least 180 days, (2) chronological age of 45 years or older, (3) presence of stroke lesion on only one side of the hemisphere (i.e., unilateral lesion), and (4) successful extraction of cortical thickness.

The collection of ENIGMA stroke data followed the Declaration of Helsinki and was approved by local ethics boards at each respective institute. All participants provide written informed consent for the study. This study was approved by the Institutional Review Board (IRB# 00002881) at the University of Southern California Health Science Campus. The UKB data used for our study was anonymous and deidentified, exempting the study from the requirement of obtaining informed consent.

### Behavioral Data

Due to the heterogeneity of motor outcomes recorded by each site in the ENIGMA stroke dataset, we harmonized different motor outcome measures (Supplementary Table 1). As done previously, we defined a primary motor outcome score based on the maximum possible score for each measure.^16,18,19^ A score of 1.0 indicates no impairment and 0.0 indicates severe impairment.

### MRI Data Processing

#### MRI scan parameters for UK Biobank

3D T1-weighted images were acquired from the UK Biobank dataset. The imaging parameters include an inversion time of 880 ms, a repetition time of 2000 ms, an echo time of 2.01 ms, a 1 mm isotropic voxel size, a matrix size of 208 x 256 x 256, and a SENSE factor of 2.^20^

#### Stroke imaging parameters

A full description about the acquisition of high-resolution T1-weighted images in the stroke dataset has been reported elsewhere.^16^ Briefly, imaging was acquired according to standardized protocols such as the Human Connectome Project or Alzheimer’s Disease Neuroimaging Initiative. Since protocols vary across sites, images were visually inspected for quality control prior to further processing, as well as after each processing step.

#### Image processing

All T1-weighted images in both the UK Biobank dataset and the ENIGMA Stroke dataset were processed using a modified CIVET pipeline to extract cortical morphometry features (http://www.bic.mni.mcgill.ca/ServicesSoftware/CIVET).^21^ The reconstructed surfaces consisted of 40,962 vertices in each hemisphere. Cortical thickness measurements were obtained by computing the Euclidean distance between the vertices of the inner cortical surface and the corresponding vertices of the outer cortical surface. The GM/WM intensity ratio was extracted based on the inner surface information.^30^ Further methodological details are provided in Supplementary Method 1.

#### Atlas generation

The cortical surface was partitioned into 9 functional subregions as defined by Yeo et al. (Fig. 2).^32^ The subregions include the sensorimotor, frontoparietal, dorsal attention, ventral attention with language, default mode, salience, auditory, visual, and limbic networks. Since we intended to study the brain age of the ipsilesional and contralesional hemispheres, the 9 functional subregions were further divided into left and right hemispheres, for a total of 18 regions of interest (ROIs). We provided details of this part in Supplementary Method 2.

**Figure 2.**
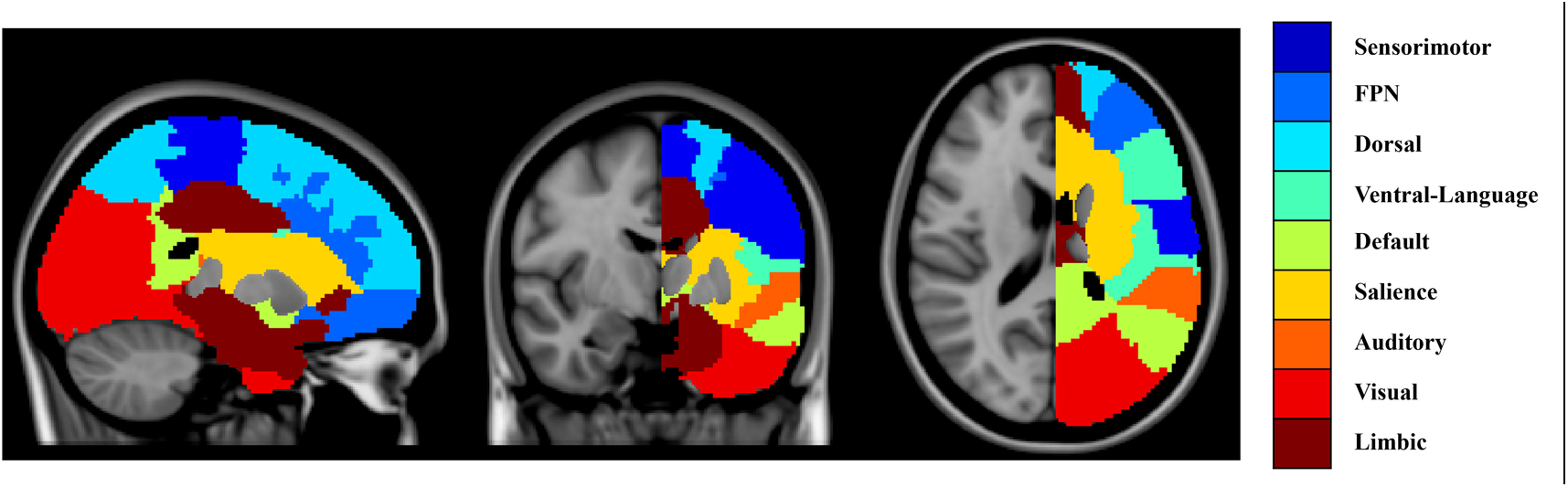
Functional subregions used to define local stroke lesion load. We extended the atlas defined by Yeo et al. (2011) to include white matter regions. Each voxel in the white matter region was labeled based on a k-nearest neighbor. The subregions of interest include the sensorimotor, frontoparietal, dorsal attention, ventral attention with language, default mode, salience, auditory, visual, and limbic networks, and these were further divided into left and right hemispheres to produce 18 regions of interest (ROIs).

### Lesion Analysis

Stroke lesions were manually segmented by trained research team members, based on a previously published protocol.^33^ Each 3D T1-weighted image and lesion mask in the stroke dataset was co-registered to MNI space using linear and nonlinear transformations generated by the CIVET pipeline as described previously.^33^

To quantify focal lesion damage for the 18 ROIs, we calculated lesion load, which is measured by dividing the volume of the overlap between the lesion and each ROI by the respective ROI volume. We also calculated a corticospinal tract (CST) lesion load (CST-LL) using a publicly available CST template that includes primary and higher order sensorimotor regions.^34^ The CST is recognized as an important predictor of motor outcome due to its importance as a motor pathway in the nervous system. ^9,34^ By including CST-LL in our model, we were able to assess the influence of local lesion loads of each ROI on motor outcomes, independent of CST-LL on motor outcome.

### Regional Brain Age Prediction

We developed an in-house regional brain age prediction model using graph convolutional networks (GCNs),^35^ trained on the UK Biobank dataset (Fig. 3). The model utilized cortical thickness and GM/WM intensity ratio at each vertex as features (Fig. 3A). Vertices and their connections were defined as nodes and edges in the graph structure, respectively Signals at each node were projected into the spectral domain via graph Fourier transform, filtered, and then projected back into the spatial domain.^36^ The GCN architecture included a graph convolutional layer, rectified linear unit activation, graph max pooling, and a fully connected layer for prediction (Fig. 3B). Five-fold cross-validation was performed, and an ensemble of the five models was used to improve the prediction performance on the target dataset. Detailed model architecture and training procedures are provided in the Supplementary Method 3.

**Figure 3.**
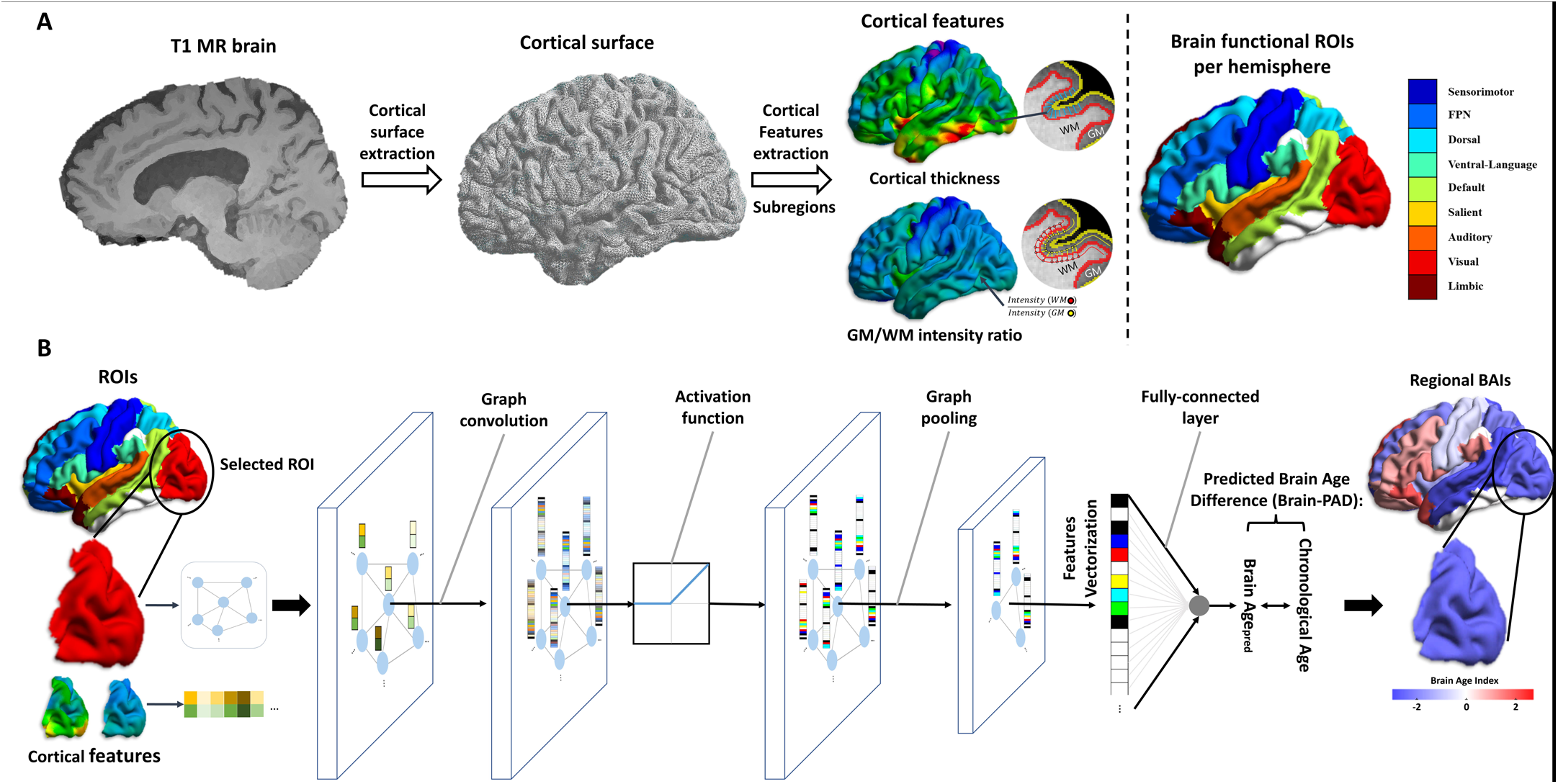
The training flowchart for predicting regional brain age. **(A)** Flowchart for data generation to pass into graph convolutional networks (GCNs) for predicting regional brain age. We built a cortical surface model from a 3D T1-weighted image using the CIVET pipeline and extracted cortical thickness and gray matter (GM) to white matter (WM) intensity ratio. The cortical surface and cortical features were divided into 18 regions of interest (ROIs). **(B)** One GCN model per ROI was trained to predict regional brain age. The cortical surface was used to define nodes and edges for a graph structure, and cortical features were used as signals for each node. All models have the same GCN structure, which consists of a graph convolution, rectified linear unit for activation function, max-pooling layer for graph pooling, and a fully connected layer. To obtain regional brain predicted age difference (brain-PAD), we took the difference between the predicted regional brain age and chronological age.

### Brain Predicted Age Difference (Brain-PAD) Analysis

Brain-PAD was calculated as the difference between the predicted brain age and the chronological age of the individual. A higher, positive brain-PAD is indicative of an older-appearing brain, whereas a lower, negative brain-PAD is indicative of a younger-appearing brain. Regional brain-PAD is defined as the brain-PAD obtained from a particular functional subregion, or ROI. To mitigate the regression dilution bias observed with brain-PAD, we employed the linear trend removal method proposed by Smith et al.^37^ This adjustment ensures that brain-PAD reflects the relative brain health status of individuals, independent of their age.

### Statistical Analysis

We first investigated the relationship between regional brain-PAD and total lesion volume. Linear mixed-effects models were employed with each regional brain-PAD as the dependent variable, total lesion volume as the independent variable, age at scan time, sex, and days since stroke as covariates and cohort as a random effect.

Next, we investigated the relationship between lesion load of each ROI, including CST-LL, and regional brain-PAD using linear mixed-effects models, with lesion load as the dependent variable, regional brain-PAD as the independent variable, and the same covariates and random effects as above. To focus on relationships between each lesion load and regional brain-PAD, we included mean regional brain-PAD as an additional covariate to account for overall brain aging patterns. For both analyses, if a lesion load was greater than 20% of the total volume of a ROI for which brain-PAD was computed, we excluded that regional brain-PAD from our analysis. This 20% cut-off was determined empirically to avoid potentially false brain age predictions due to lesions encroaching on the region. False discovery rate (FDR) correction was applied to account for multiple comparisons.

We then predicted motor outcomes using different machine learning methods: random forest,^39^ gradient boosting,^40^ AdaBoost,^41^ and XGBoost.^42^ Based on previous literatures,^9,38^ A threshold of 0.636 was used to categorize motor outcomes into good (mild to no impairment; n = 278) and poor (moderate to severe impairment; n = 223). Performance metrics such as accuracy and area under the curve (AUC) were used to evaluate the predictive quality of each method. Input features for these methods included local lesion loads, CST-LL, regional brain-PAD, age at scan time, sex, days since stroke, and ICV. The goal of this prediction was to determine the impact of each feature on motor outcomes. To do this, we calculated the Gini index, which estimates the impurity, or the probability of incorrectly labeling a randomly chosen element in the dataset, at each node of the decision tree. A Gini index of 0 suggests complete purity (i.e., only one label in the set), whereas a Gini index of 0.5 suggests complete impurity (i.e., equally likely to obtain both labels in the set). To establish feature importance, we determined how much each feature increased the purity by decreasing the Gini index. In other words, the features with the highest importance contributed to the greatest magnitude decrease in the Gini index, which we refer to as the feature importance score. To derive a statistically robust importance score, we performed bootstrapping with 5,000 iterations where the dataset was randomly divided into training and test sets with 8:2 splits. We examined the mean and standard deviation of the feature importance scores obtained in the 5000-iteration bootstrap.

In our final analysis, we examined directionality within the relationships between CST-LLs, regional brain-PADs, and motor scores using structural equation modeling (SEM). Based on our hypotheses, we evaluated the mediating effects of ipsilesional regional brain-PAD and motor impairment severity on mean contralesional regional brain-PAD. Models were fitted to all data by an optimizer with a Wishart log likelihood, and model fit was assessed using a chi-squared distribution, comparative fit index (CFI), root mean square error of approximation (RMSEA), and adjusted goodness-of-fit index (AGFI). Implementation details for all models are available in Supplementary Method 4.

## Results

As of June 23, 2023, the ENIGMA Stroke Recovery Working Group dataset contained data from 883 patients who matched our initial eligibility criteria. Cross-sectional data from 501 individuals with stroke from 34 cohorts across 10 countries met the final eligibility criteria and were included in this analysis. There were 318 male, 162 female, and 21 unknown sex patients, with a median age of 63 years (interquartile range [IQR] 14 years). Stroke severity (i.e., primary sensorimotor score) ranged from 0 to 1 (median 0.6516, IQR 0.4916). Days after stroke ranged from 180 to 8,915 days (median 704 days, IQR 1,259 days). Lesion volume ranged from 0.013 to 339.64 mL (median 7.35 mL, IQR 49.04 mL) and a lesion overlap map was produced (Supplemental Fig. 1).

Using the UK Biobank dataset, we assessed the performance of the brain age prediction models for each region of interest (ROI). The mean absolute errors (MAE) ranged from 2.94 to 3.13 years, and correlation coefficients (R-values) ranged from 0.88 to 0.90, indicating robust predictive performance across all models (Table 1).

**Table 1.** Regional brain age prediction results of the UKB dataset. The mean absolute error (MAE) and correlation coefficient (R-value) between predicted age and chronological age for the brain age prediction model for each region of interest are shown. MAEs range from 2.94 to 3.13, and R-values range from 0.88 to 0.90. All models demonstrate similar robust performance in predicting brain age.

|  |  | Sensorimotor | FPN | Dorsal | Ven-Lan | Default | Salience | Auditory | Visual | Limbic |
| --- | --- | --- | --- | --- | --- | --- | --- | --- | --- | --- |
| Left | MAE | 3.01 | 3.01 | 2.94 | 3.05 | 3.08 | 3.04 | 2.99 | 3.06 | 3.08 |
|  | R-value | 0.89 | 0.89 | 0.90 | 0.89 | 0.89 | 0.89 | 0.89 | 0.88 | 0.89 |
| Right | MAE | 3.13 | 2.94 | 2.94 | 2.98 | 3.09 | 3.05 | 3.07 | 3.12 | 3.05 |
|  | R-value | 0.88 | 0.90 | 0.90 | 0.89 | 0.89 | 0.89 | 0.89 | 0.88 | 0.89 |

### Larger total stroke lesion size is positively correlated with higher ipsilateral regional brain-PADs

We first examined the relationship between total stroke lesion size and regional brain-PAD of the 18 functional ROIs (Table 2). Larger total lesion size was positively correlated with regional brain-PADs for all ipsilesional ROIs except for the auditory and visual networks (FDR-corrected p-value < 0.05). Larger total lesion size was negatively correlated with brain-PAD in the contralesional ventral attention-language network ROI (FDR-corrected p-value < 0.05). No significant associations were observed between total lesion size and other contralesional ROIs.

**Table 2.**
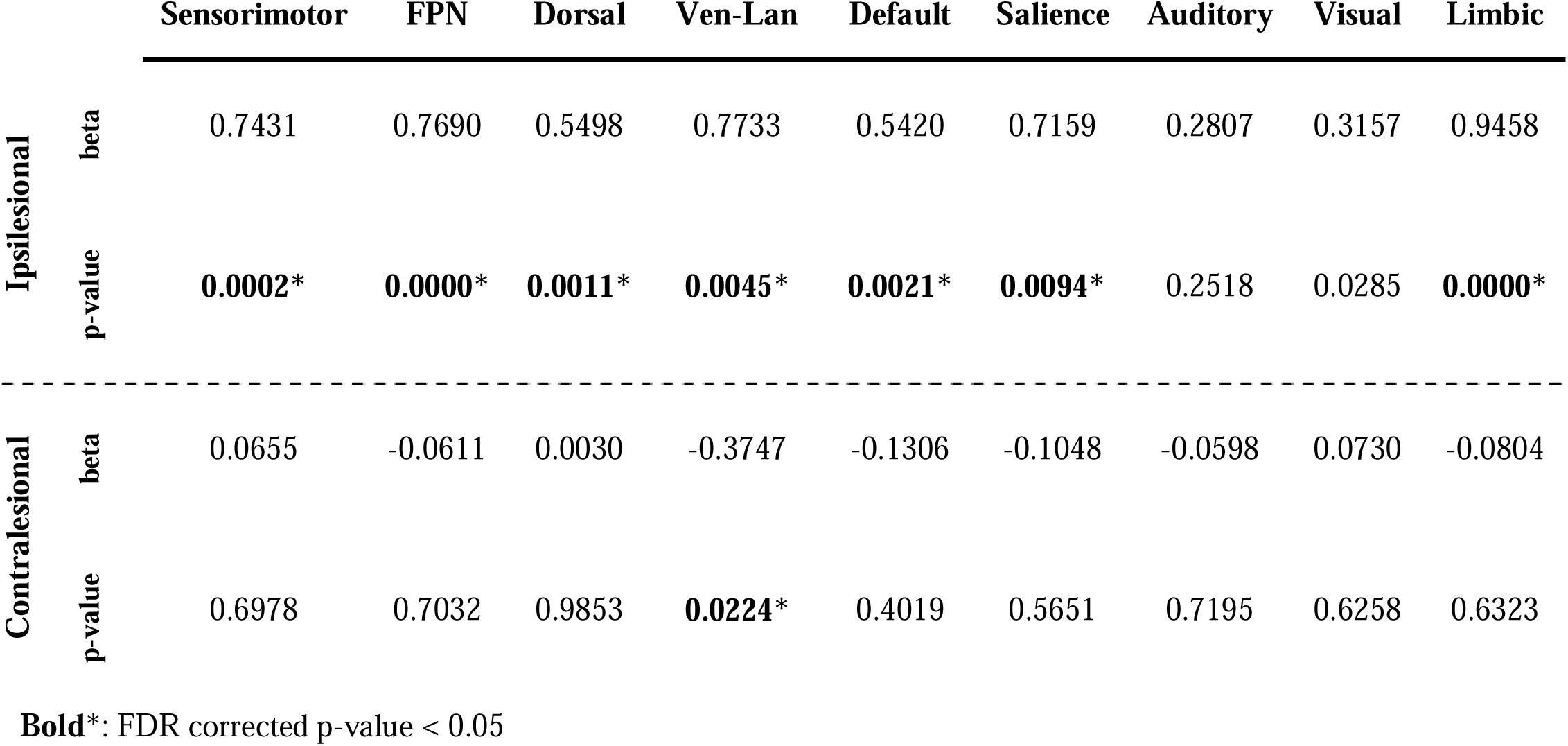
Larger total stroke lesion size is correlated with higher ipsilesional regional brain-PAD. The beta coefficient and significance of the effect of total stroke lesion size on each region of interest after applying FDR correction is shown. A higher total stroke lesion size was significantly correlated with higher regional brain-PAD in all ipsilesional regions except the auditory network and visual network and lower regional brain-PAD in only the contralesional ventral attention-language network.

### Higher local lesion load is correlated with higher ipsilesional and lower contralesional regional brain-PADs

The second analysis examined the relationship between regional brain-PAD and local lesion loads of the 18 functional networks, as well as the CST-LL (Fig. 4). Higher local lesion loads were correlated with higher ipsilesional regional brain-PAD and lower contralesional regional brain-PAD. Regional brain-PADs of the ipsilesional FPN, default mode, salience, and visual networks, as well as the contralesional FPN, dorsal attention, ventral attention-language, and salience networks were associated with at least three lesion load metrics each. Notably, higher lesion loads in the salience network showed widespread significant associations in regional brain-PADs across both ipsilesional (sensorimotor, FPN, ventral attention-language, default mode, and visual networks) and contralesional (FPN, dorsal attention, ventral attention-language, default mode, salience, and auditory networks) hemispheres.

**Figure 4.**
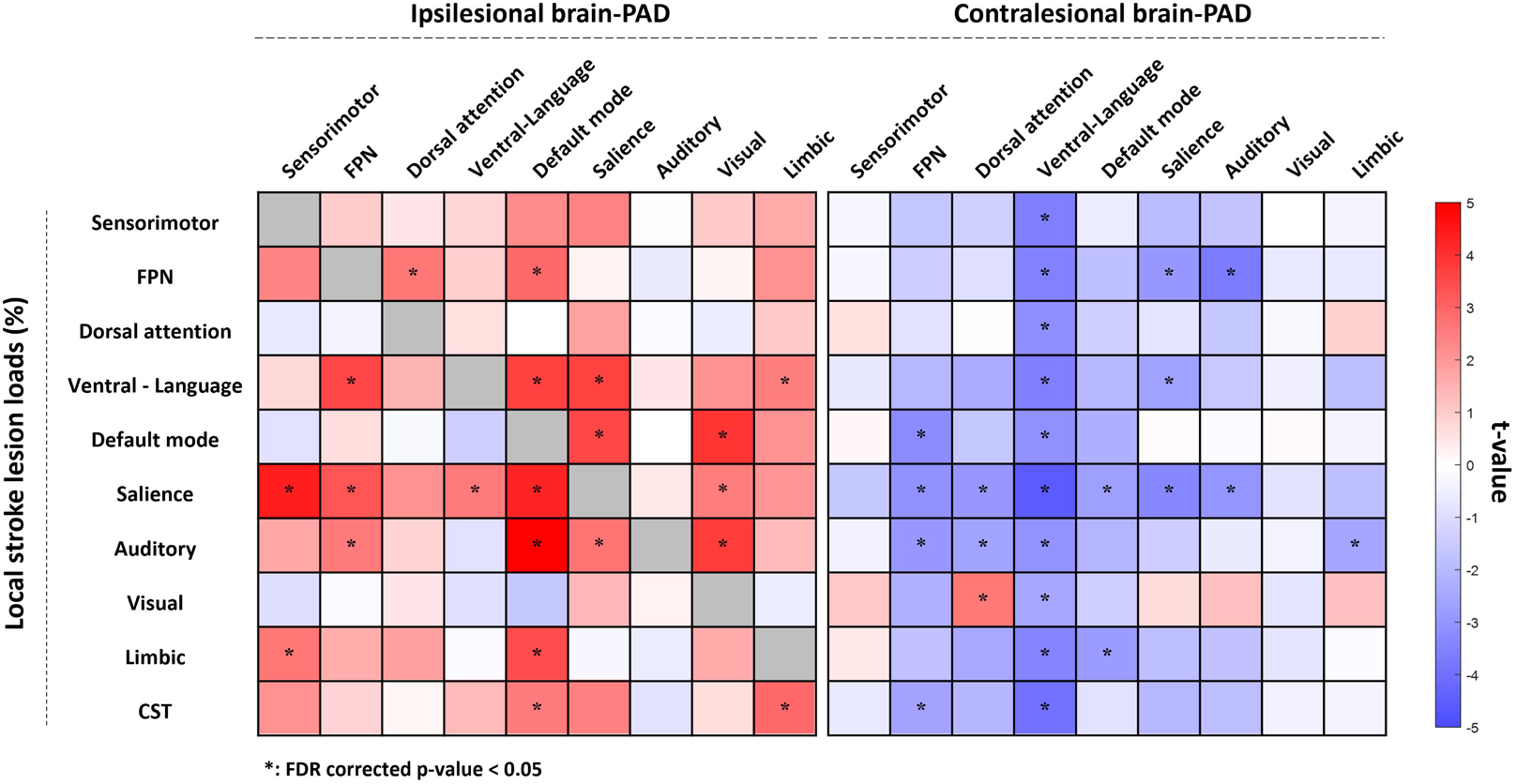
Results of the association analysis between lesion loads and regional brain-PAD. Each square represents the association between regional brain-PAD (x-axis) and lesion load (y-axis). A darker red color indicates higher brain-PAD (older appearing brain), while a darker blue color indicates lower brain-PAD (younger appearing brain). As the brain age in lesional regions (i.e. a lesion load in the ROI for which brain-PAD was computed > 20%) was not computed due to a potentially false brain age prediction, the correlation analyses in the diagonal cells were not performed and colored in gray. The asterisk denotes a significant result (p < 0.05) after False Discovery Rate (FDR) correction. Higher local stroke lesion loads showed correlations with higher ipsilesional and lower contralesional regional brain-PADs. Specifically, ipsilesional regional brain-PADs of FPN, default mode, salience, and visual networks and contralesional regional brain-PADs of FPN, dorsal attention, ventral attention-language, and salience networks were influenced by at least 3 lesion loads. Lesion load in the salience network showed widespread significant influence on both ipsilesional (sensorimotor, FPN, ventral attention-language, default mode, and visual networks) and contralesional regional brain-PAD (FPN, dorsal attention, ventral attention-language, default mode, salience, and auditory networks).

### Regional brain-PAD of contralesional networks and local lesion loads have the highest importance in predicting motor outcome

We evaluated features important for predicting motor outcome. The random forest method showed the most robust performance in terms of both accuracy (median: 0.6634) and AUC (median: 0.6572) and was used to evaluate the importance of all input features (Supplementary Table 2). The top three predictive features based on mean importance scores were CST-LL (mean importance score [MIS]□=□0.0646), salience network lesion load (MIS□=□0.0593), and contralesional FPN brain-PAD (MIS□=□0.0491) (Figure 5). Importantly, only contralesional—and not ipsilesional—regional brain-PADs were significant predictors of motor outcomes, with younger contralesional brain-PAD associated with worse motor impairment.

**Figure 5.**
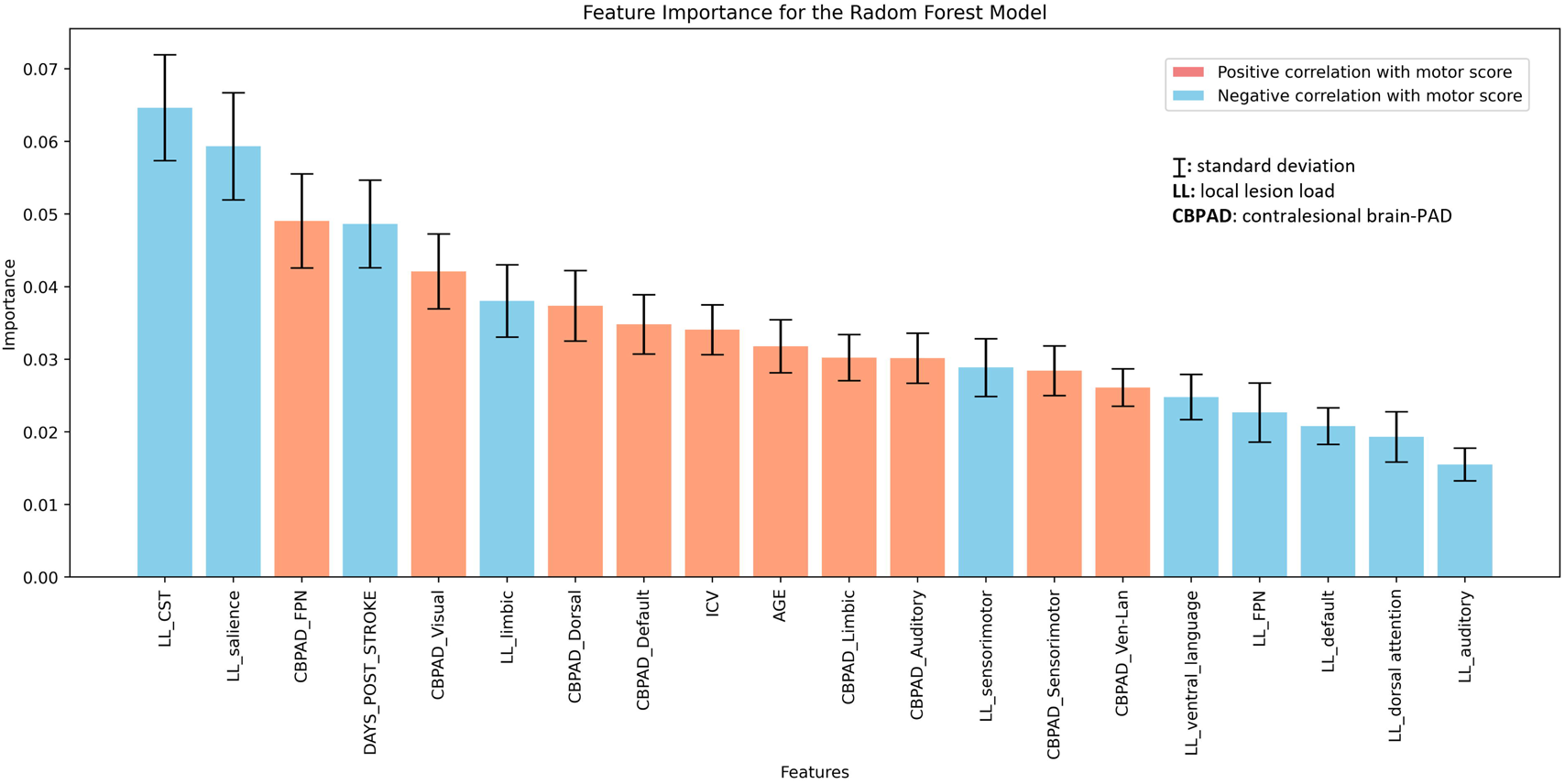
Ranking of feature importance scores in predicting motor outcome based on 5,000 iterations bootstrapping of the Random Forest method. The mean and standard deviation of the feature importance scores are shown for each predictor of motor outcome. The color of the bars indicates whether each feature is significantly correlated with a motor score value, indicating a positive (light red) or negative (light blue) trend. The results highlight the top three features based on mean importance: local lesion load in the corticospinal tract and salience network, and contralesional regional brain-PAD in the FPN. Of the top 20 significant predictors, 8 were contralesional regional brain-PAD and none were ipsilesional regional brain-PAD.

### Motor outcomes mediate the impact of CST-LL on mean contralesional regional brain-PAD

We employed structural equation modeling (SEM) to examine the directional relationships among CST-LL, motor outcomes, mean contralesional brain-PAD, and ipsilesional brain-PAD (Fig. 6). The SEM model had acceptable model fit indices: χ^2^/df=0.99, χ^2^ p-value = 0.3717, CFI=1.00, RMSEA=0.00, and AGFI=0.95.^43,44,45^ Higher CST-LL was directly associated with worse motor outcomes (β□=□–0.355, p□<□0.001) and higher ipsilesional brain-PAD (β□=□0.262, p□<□0.001). Elevated ipsilesional brain-PAD was directly associated with worse motor outcomes (β□=□–0.102, p□<□0.05) and higher contralesional brain-PAD (older brain aging; β□=□0.213, p□<□0.001). Worse motor outcomes were directly associated with lower contralesional brain-PAD (younger brain aging; β = 0.204, p-value < 0.001). These results suggest that motor outcome mediate the impact of CST-LL on the contralesional brain-PAD. Higher CST-LL contributed to lower contralesional brain-PAD via its association with motor outcomes (indirect effect□=□– 0.072), whereas higher CST-LL was associated with lower motor outcome scores through its effect on ipsilesional brain-PAD (indirect effect = −0.028).

**Figure 6.**
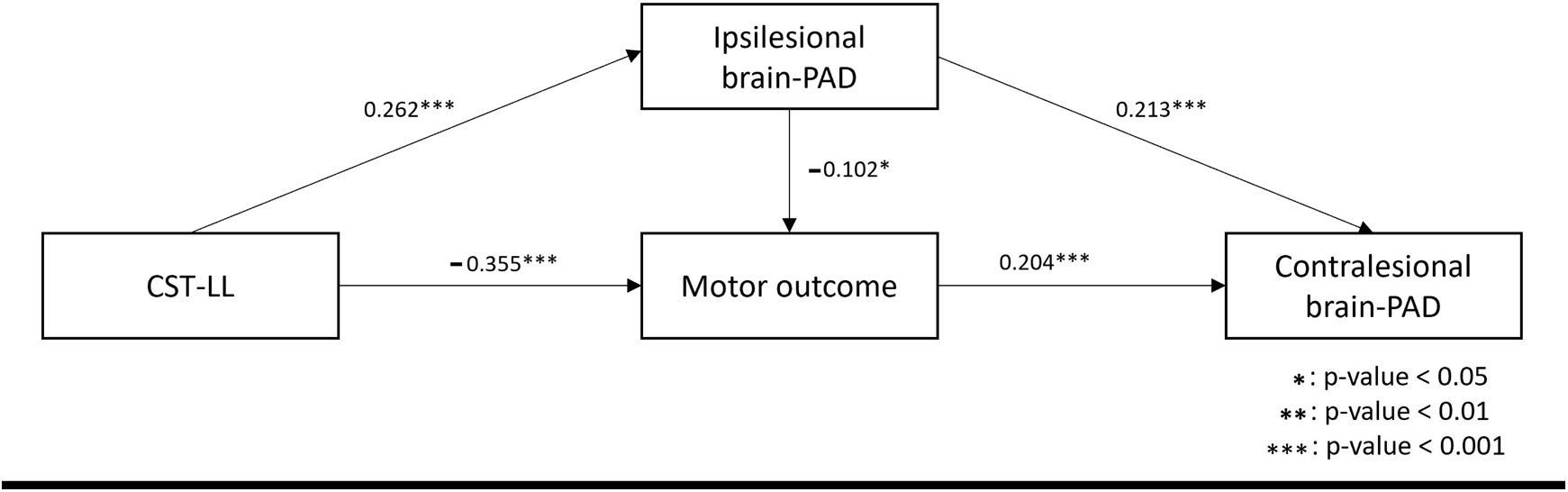
Structural equation model (SEM) to determine the relationship between CST- LLs, regional brain-PADs, and motor scores. This model shows the significant directional relationship between corticospinal tract lesion load (CST-LL), ipsilesional brain-PAD, mean contralesional regional brain-PAD, and motor outcome. The SEM model had acceptable model fit statistics: χ2/df=0.99, χ2 p-value = 0.3717, CFI=1.00, RMSEA=0.00, and AGFI=0.95. All associations in the model were statistically significant (p-value < 0.001) except for the association between ipsilesional brain-PAD and motor outcome. A higher CST-LL correlated with worse motor outcomes (β = −0.355, p-value < 0.001) and higher ipsilateral brain-PAD (β = 0.262, p-value < 0.001). A higher ipsilesional brain-PAD correlated with worse motor outcome (β = −0.102, p-value < 0.05) and higher mean regional brain-PAD in contralesional networks (β = 0.213, p-value < 0.001). A worse motor outcome correlated with lower mean regional brain-PAD of contralesional networks (β = 0.204, p- value < 0.001). We also found that motor outcome mediated the impact of CST-LL on mean regional brain-PAD of contralesional networks. More specifically, CST-LL directly and negatively affected motor outcomes (direct effect=-0.355), with larger lesion load associated with worsened motor impairment. CST-LL indirectly and negatively affected motor outcomes through its effect on ipsilesional brain-PAD (indirect effect=-0.028). On the other hand, CST-LL indirectly and negatively affected mean regional brain-PAD of contralesional networks through its effect on motor outcomes (indirect effect=-0.072). Ipsilesional brain-PAD directly and positively affected the mean regional brain-PAD of contralesional networks (direct effect=0.213).

## Discussion

In this study, we demonstrate a complex relationship between total and focal lesion damage, regional brain age measures, and motor outcomes in patients with chronic unilateral stroke. We found that larger total lesion sizes and higher local lesion loads were associated with higher ipsilesional brain-PADs and lower contralesional brain-PADs. Notably, lesion load in the salience network significantly influenced regional brain-PADs across both hemispheres. Additionally, CST-LL, lesion load in the salience network, and regional brain-PAD in the contralesional FPN network emerged as the most significant predictors of motor outcomes. We also found that higher CST-LL was associated with older ipsilesional brain-PADs and worse motor impairment, consistent with a previous study^9^. Interestingly, more severe motor impairment was associated with younger contralesional brain-PADs, suggesting that motor impairment may drive compensatory changes in the contralesional hemisphere.

Our regional brain age model confirmed our hypothesis that in unilateral chronic stroke, larger stroke volumes and higher lesion loads were associated with higher brain-PADs in ipsilesional brain regions. Conversely, higher lesion loads were associated with lower brain-PADs in contralesional regions. Since mean regional brain age was included in the model, these findings indicate region-specific patterns independent of overall brain aging patterns. This shows that regional brain age is sensitive to focal and network-specific damage following stroke in ways that global brain age may not fully capture.

Lesion overlap with critical brain regions are one of the most important features in predicting motor outcomes after stroke. The most important feature in our prediction models was the CST-LL, as expected based on prior literature.^9,34^ Interestingly, the second most important feature was lesion load of the salience network. Lesion load in the salience network was strongly associated with older ipsilesional brain-PADs and younger contralesional brain-PADs in almost all regions. Damage to the salience network disrupts connections with other functional networks like the default mode network and FPN.^47^ Given that imbalances in the salience network have been linked to cognitive and neuropsychiatric disorders,^48^ future studies could assess relationships between damage to the salience network, cognitive and emotional dysfunction, and motor impairment following stroke.

Following the lesion load features, the next most important feature to predict motor outcome was younger contralesional brain-PAD in the FPN. The FPN modulates activity of other brain networks like the default mode network, thus playing an important role in cognition and motor control.^49^ Importantly, increased activity and connectivity in contralesional FPN structures have been observed during the subacute and chronic stages of stroke, promoting recovery of motor function.^50,51^ Further studies could elucidate whether the FPN is a strong target for neuromodulation to improve motor outcomes following stroke. Intriguingly, among the 20 significant motor outcome predictors, eight were younger contralesional brain-PADs, and none were ipsilesional brain-PADs, suggesting that decelerated aging in the contralesional hemisphere may occur in response to severe motor impairment. One explanation is increased reliance on the non-paretic side, leading to enhanced connectivity in the contralesional hemisphere controlling it.^52^ Such changes might manifest as a younger brain-PAD in our study.^53^ Alternatively, severe impairment may necessitate more widespread brain activation in both hemispheres during paretic limb use.^12^ This recruitment may occur via the corpus callosum,^54,55,56^ may compensate for damaged ipsilesional tissue. Future studies should use longitudinal designs to investigate the time course of regional brain age changes and their causal relationship with motor outcomes.

In the absence of longitudinal data, we used structural equation modeling (SEM) with cross-sectional data to investigate possible directional relationships between regional brain changes and motor outcomes. We found that motor impairment mediated the relationship between CST damage, ipsilesional brain age, and regional brain age in the contralesional hemisphere. CST damage and increased brain age are thought to be measures of primary (focal) damage and secondary atrophy due to stroke, respectively; as CST damage and ipsilesional brain age increased, we observed worse motor outcomes. In turn, we found that worse motor outcomes result in younger mean contralesional regional brain age, which is consistent with our hypotheses and may suggest that the brain utilizes contralesional regions to compensate for severe motor impairment. Importantly, these findings demonstrate the complex, bidirectional nature of the relationship between brain aging and motor impairment after stroke, as ipsilesional brain age impacts motor outcomes, which then drive changes in contralesional brain age.

A limitation of this work is its cross-sectional design and focus on the chronic phase (>180 days post-stroke). Future research should examine brain age from acute stages onward to understand when the relationship between contralesional brain age and motor outcomes is strongest, potentially identifying optimal windows for intervention. Additionally, investigating the relationship between contralesional brain age and non-paretic limb usage, possibly using wearable sensors, could enhance our understanding of how behavioral adaptations influence regional brain aging.

Our findings emphasize the importance of assessing regional brain age as a sensitive biomarker for neuroplasticity and motor recovery after stroke. The observed accelerated aging in ipsilesional regions and paradoxically younger brain age in contralesional regions suggest different roles for each hemisphere in response to stroke-induced damage. Targeting specific neural networks, such as the contralesional FPN and salience network, may hold promise for developing personalized rehabilitation strategies aimed at enhancing motor recovery.

## Data Availability

UK Biobank data is available by request to the UK Biobank group (https://www.ukbiobank.ac.uk/enable-your-research/apply-for-access). ENIGMA Stroke Recovery data is available by request to the ENIGMA Stroke Recovery group (https://enigma.ini.usc.edu/ongoing/enigma-stroke-recovery/), subject to local PI approval and compliance with all relevant regulatory boards.

## Acknowledgements

G. Park was supported by Micheal J Fox Foundation (MJFF) - The Parkinson’s Progression Markers Initiative (PPMI) #MJFF-023385. M. Khan and S.-L. Liew were supported by the National Institutes of Health (NIH) grant R01 NS115845. L. Boyd was supported by Canadian Institutes of Health Research (CIHR) Operating grants (MOP-130269; MOP- 106651) and Project grant (PTJ-148535) PI L.A.B. A. Brodtmann was supported by a National Health and Medical Research Council project grant GNT1020526, the Australian Brain Foundation, Wicking Trust, Collie Trust, and Sidney and Fiona Myer Family Foundation, as well as fellowships from the National Heart Foundation 100784 and 104748. C. Buetefisch was supported by NIH grant (R01NS090677). A. B. Conforto was supported by Hospital Israelita Albert Einstein 2250-14, and NIH R01NS076348-01. N. Egorova-Brumley was supported by the Australian Research Council Future Fellowship FT230100235. F. Geranmayeh was supported by the Wellcome Trust (093957), and partly supported by the National Institute for Health Research (NIHR) Imperial Biomedical Research Centre. C. A. Hanlon was supported by NIH/National Institute of General Medical Sciences (NIGMS) grant (P20GM109040). S. Kautz was supported by NIH grant GM109040. K. Revill was supported by NIH grant (R01NS090677). N. J. Seo was supported by grants from the NIH/National Institute of Child Health and Human Development (NICHD) (R01HD094731) and NIH/NIGMS (P20GM109040). S. Soekadar was supported by European Research Council (ERC NGBMI 759370), Deutsche Forschungsgemeinschaft (DFG) grant SO 932/7-1. G. Thielman was supported by the REACT Pilot Grant sub-award for “The effect of transcranial direct current stimulation (tDCS) on upper extremity use following a Cerebral Vascular Accident (CVA)” and the National Resource Center for High-Impact Clinical Trials in Medical Rehabilitation: Pilot Projects Component 2019-2021, Primary Investigator. D. Vecchio was supported by RC-24. L. T. Westlye was supported by the Research Council of Norway [249795, 248238], the South-Eastern Norway Regional Health Authority [2014097, 2015044, 2015073, 2018037, 2018076, 2019107, 2020086], the Norwegian Extra Foundation for Health and Rehabilitation [2015/ FO5146], Sunnaas Rehabilitation Hospital HT, and the Department of Psychology, University of Oslo. L.T. Westlye performed this work on the Services for sensitive data (TSD), University of Oslo, Norway, with resources provided by UNINETT Sigma2 - the National Infrastructure for High-Performance Computing and Data Storage in Norway. G. F. Wittenberg was supported by grants from the VA Rehabilitation Research and Development program and the NIH/National Institute of Neurological Disorders and Stroke (NINDS). H. Kim was supported by NIH grant (#U01 AG024904).

