## Supplemental appendix for "Severe motor impairment is associated with lower contralesional brain age in chronic stroke"

### Supplementary appendix

**Supplementary Method 1. MRI Data Processing: Image processing**

All T1-weighted images in both the UK Biobank dataset and the ENIGMA Stroke dataset were processed using a modified CIVET pipeline to extract cortical morphometry features (<http://www.bic.mni.mcgill.ca/ServicesSoftware/CIVET>).^1^ The pipeline includes non-uniform intensity correction,^2^ skull stripping,^3^ registration to a stereotaxic space,^4^ and brain tissue segmentation.^5,6^ Based on the brain tissue information, inner and outer cortical surface models were reconstructed using Constrained Laplacian-Based Automated Segmentation with Proximities algorithm.^7^ The reconstructed surfaces consisted of 40,962 vertices in each hemisphere. The cortical surface models were inversely registered into the native space,^8^ a process that employed an iterative surface registration to guarantee an optimal correspondence at each vertex across individuals.^9^ Cortical thickness measurements were obtained by computing the Euclidean distance between the vertices of the inner cortical surface and the corresponding vertices of the outer cortical surface. The GM/WM intensity ratio was extracted based on the inner surface information.^10^ In the stroke dataset, constructing the cortical surface model can be challenging due to the presence of stroke regions. To alleviate this problem, we employed a method of filling the stroke region by mirroring the corresponding region in the contralesional hemisphere.^11^ Empirically, this method reduced the number of failed cases by 35% compared to the method using raw images.

**Supplementary Method 2. MRI Data Processing: Atlas generation**

The cortical surface was partitioned into 9 functional subregions as defined by Yeo et al. (Fig. 2).^12^ The subregions include the sensorimotor, frontoparietal, dorsal attention, ventral attention with language, default mode, salience, auditory, visual, and limbic networks. We used the Automated Anatomical Labeling (AAL) template to provide anatomical parcellation in the MNI space. As the atlas contained only gray matter voxels, we expanded the atlas to include neighboring white matter regions using a k-nearest neighbor approach. The complete gray and white matter atlas were generated in MNI space, using the fitcknn function in MATLAB 2022b with a parameter k value of 50 and the Minkowski distance metric. Since we intended to study the brain age of the ipsilesional and contralesional hemispheres, the 9 functional subregions were further divided into left and right hemispheres, for a total of 18 regions of interest (ROIs).

**Supplementary Method 3. Regional Brain Age Prediction**

An in-house regional brain age prediction model was developed and trained on the UK Biobank dataset using CIVET-extracted morphological features of cortical thickness and GM/WM intensity ratio at each vertex. This model was then used to predict regional brain ages in the stroke patient dataset.

To extract regional brain ages from each of the 18 ROIs noted above, we used graph convolutional networks (GCNs),^13^ exploiting the data with graph structure and signal at each node (Fig. 3). A vertex in the cortical surface model and connectivity between vertices were defined as the node and edge for the graph structure, respectively. Cortical features (thickness and GM/WM intensity ratio) were used as signals at each node. In GCNs, the signal–as a feature vector of size 2–was projected into the spectral domain by the graph Fourier transform based on the eigenvectors of the normalized graph Laplacian.^14^ It was filtered by weights with learnable parameters and projected into the original domain by a graph Fourier inverse transform. The filtered features underwent an activation function for nonlinearity and a graph pooling to aggregate features at each node. To ensure efficient pooling operations without any loss of information, a balanced binary tree was constructed from the coarsest to the finest level by introducing fake nodes or disconnected nodes. In this study, GCNs were constructed for each subregional surface mesh. The GCN architecture contained a graph convolutional layer, a rectified linear unit (ReLU) activation function, a graph max pooling operation, and a fully connected layer for regional brain age prediction. Fig. 3B shows the overall flow of our GCN model. Other details of the training process included: mean square error as the loss function, Adam optimizer, 800 epochs, a learning rate of 10e-6, L2 regularization with a weight of 10e-4 to prevent overfitting, and a batch size of 2.

T1-weighted images acquired from the UK Biobank assessment sites were used to train the regional brain age algorithm. We performed a 5-fold cross-validation using the UKB dataset to train the regional brain age models. After cross-validation, we obtained five trained models, each with its own set of learned parameters. We created an ensemble of these five models from the 5 folds to leverage collective knowledge and improve prediction performance.

**Supplementary Method 4. Statistical Analysis**

The linear mixed-effects models were implemented using the *fitlme* function in MATLAB 2022b. The random forest, gradient boosting, and AdaBoost models were implemented using Python package *scikit-learn* (v1.0.2), while the XGBoost model was implemented using Python package *XGBoost* (v1.6.2). Structural equation modeling was implemented using the Python package semopy.^15^ The hyperparameters for all methods followed default configurations.

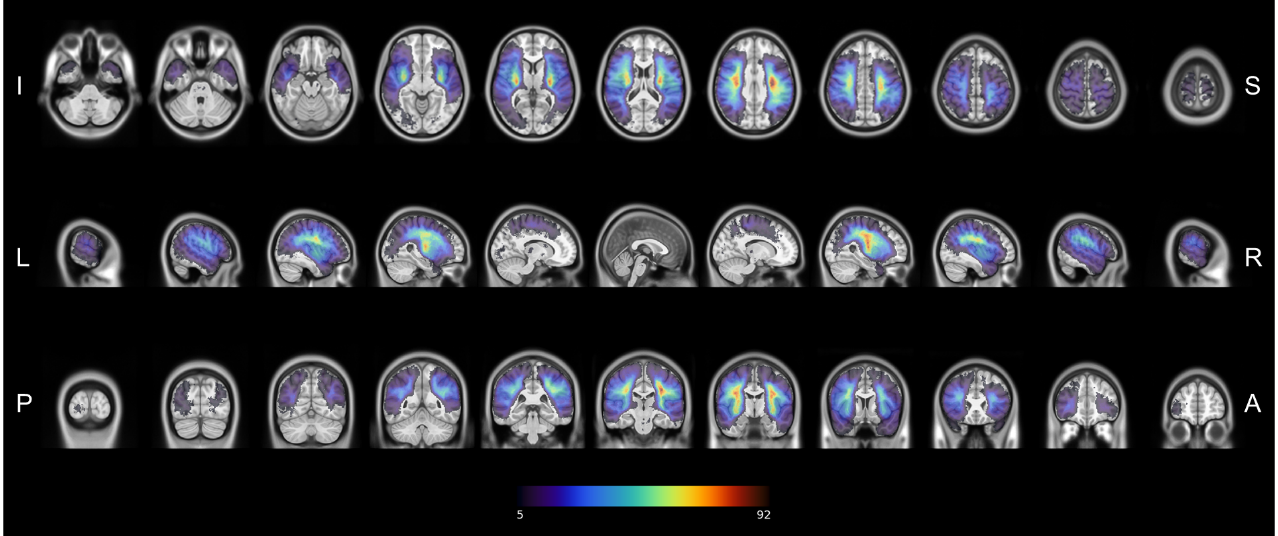

**Supplementary Figure 1. Lesion overlap maps for the individuals with chronic stroke superimposed on the MNI brain template.** Slices are shown in the axial, sagittal, and coronal plane. The color scale indicates the number of subjects with a lesion at each voxel. The voxel with maximum overlap consists of 92 individuals. Inferior (I) to Superior (S), Left (L) to Right (R), and Anterior (A) to Posterior (P) orientations are indicated.

| **Behavioral Measures** | **Total Subjects** | **Sites** | **Possible Score Range** |
| --- | --- | --- | --- |
| Fugl Meyer Upper Extremity | 421 | R001, R002, R003, R004, R005, R010, R011, R015, R017, R018, R023, R027, R028, R029, R030, R031, R033, R034, R035, R042, R044, R045, R046, R047, R048, R052, R057 | 0-66 |
| Motor Component of the Cognitive Assessment at Bedside for iPad (CABPAD) | 34 | R009 | 0-55 |
| Medical Research Council Scale for Muscle Strength | 19 | R024 | 0-5 |
| Normalized Grip Strength | 9 | R021, R025 | 0-1 |
| Burden of Stroke Scale | 8 | R022 | 0-100 |
| Peak VO_2_ Normalized for Age and Sex | 7 | R025 | 0-100 |
| NIH Stroke Scale | 3 | R041, R053 | 0-42 |

**Supplementary Table 1: Harmonization of behavioral measures to derive a primary sensorimotor outcome score.**

Each measure was selected to reflect sensorimotor outcomes. The subsequently-derived primary sensorimotor score is a continuous measure where 1.0 indicates no impairment and 0.0 indicates severe impairment. The sites and total number of subjects with each measure are also indicated.

**Supplementary Table 2. Performance metrics of predicting motor outcome for each machine learning model.**

|  | Random Forest | Gradient Boosting | AdaBoost | XGBoost |
| --- | --- | --- | --- | --- |
| Accuracy (median) | **0.6634*** | 0.6535 | 0.6337 | 0.6535 |
| Accuracy (top 5%) | **0.7519*** | 0.7426 | 0.7175 | 0.7371 |
| Accuracy (max) | **0.8119*** | 0.8020 | 0.7921 | 0.7921 |
| AUC (median) | **0.6572*** | 0.6497 | 0.6228 | 0.6445 |
| AUC (top 5%) | **0.7481*** | 0.7357 | 0.7128 | 0.7326 |
| AUC (max) | **0.8117*** | 0.7888 | 0.7913 | 0.7895 |

**Bold***: best performance

The median, top 5%, and maximum accuracy and area under the curve (AUC) metrics are shown for each of the four machine learning models used to predict motor outcome as good or poor. Features used for prediction included lesion loads, regional brain-PADs, age, sex, days since stroke, and intracranial volume. Random forest showed the most robust performance. The asterisk denotes significantly different from other methods (p<0.05, ANOVA test).
